# The REinfection in COVID-19 Estimation of Risk (RECOVER) study: Reinfection and serology dynamics in a cohort of Canadian healthcare workers

**DOI:** 10.1101/2022.02.10.22269967

**Authors:** Étienne Racine, Guy Boivin, Yves Longtin, Deirde McCormack, Hélène Decaluwe, Patrice Savard, Matthew P. Cheng, Marie-Ève Hamelin, Fazia Tadount, Kelsey Adams, Benoîte Bourdin, Sabryna Nantel, Vladimir Gilca, Jacques Corbeil, Gaston De Serres, Caroline Quach

## Abstract

**Background:** Understanding the immune response to natural infection by SARS-CoV-2 is key to pandemic management, especially in the current context of emerging variants. Uncertainty remains regarding the efficacy and duration of natural immunity against reinfection.

**Method:** We conducted an observational prospective cohort study in Canadian healthcare workers (HCWs) with a history of PCR-confirmed SARS-CoV-2 infection to : (i) measure the average incidence rate of reinfection and (ii), describe the serological immune response to the primary infection.

**Results:** We detected 5 cases of reinfection over 14 months of follow-up, for a reinfection incidence rate of 3.3 per 100 person-years. Median duration of seropositivity was 420 days in symptomatics at primary infection compared to 213 days in asymptomatics (p<0.0001). Other variables associated with prolonged seropositivity for IgG against the spike protein included age 55 and above, obesity, and non-Caucasian ethnicity.

**Summary:** Among healthcare workers, the incidence of reinfection with SARS-CoV-2 following a primary infection remained rare, although our analysis predates the circulation of the Omicron variant.

## INTRODUCTION

Since its appearance in Wuhan (China) in December 2019, the severe acute respiratory syndrome coronavirus 2 (SARS-CoV-2) that causes coronavirus disease (COVID-19) spread into a global pandemic, leading to more than 340 million reported cases and over 5.5 million confirmed deaths as of January 24^th^ 2022, according to the WHO [1]. COVID-19 continues to exert a high burden on healthcare systems across the world because of effective interhuman transmissibility and clinical illness that leads to hospitalization in severe cases. To curb virus transmission and avoid overwhelming healthcare systems, different non-pharmacological interventions (NPIs) have been implemented, including physical/social distancing, improved hand hygiene adherence, mask mandates, business and school closures, city-wide lockdowns, and international border closures. While these mitigation measures have caused significant economic, social, and health-related adverse effects [2], they remain necessary until a sufficient proportion of individuals become protected against SARS-CoV-2 infection. With the recent emergence of the highly transmissible Delta and Omicron variants, this proportion could be higher than 90% [3].

Protection against SARS-CoV-2 infection may be acquired by recovering from a previous episode of natural infection. However, the duration of natural immunity is unknown and infection of a large proportion of the population may not suffice to achieve collective immunity, in particular when facing emerging variants. This is of concern because of unequal vaccine distribution across the world [4] and significant level of vaccine hesitancy in Europe and the United States [5]. Therefore, it is important to determine if individuals with a history of PCR-confirmed SARS-CoV-2 infection are protected against symptomatic reinfection and viral shedding and if so, how long this protection lasts.

Several large-scale prospective and retrospective cohort studies have recently addressed SARS-CoV-2 reinfection epidemiology in both healthcare workers [6–9] and general populations [10–16]. All these studies report that reinfections are generally uncommon events (less than 1% risk over several months following primary infection) and that a history of previous infection confers protection against future infection (ranging from 82% to 93%). This protection persists for at least a few months, but its long-term duration remains largely unknown. Moreover, investigations are warranted to determine if vaccination alters the acquired natural immunity against SARS-CoV-2. Recent evidence suggests that the risk of reinfection could be significantly higher with the new Omicron variant compared to previous variants [17].

The primary objective of the REinfection in COVID-19 Estimation of Risk (RECOVER) study is to estimate the incidence rate of reinfection with SARS-CoV-2 in a population of healthcare workers (HCWs) with a history of PCR-confirmed SARS-CoV-2 infection, acquired during the first or second wave of the pandemic. We describe in detail all cases of reinfection detected during the first year of the study, estimate the reinfection incidence rate in unvaccinated HCWs, describe the serological response following infection and provide an in-depth profile of a cohort of previously infected HCWs.

## METHODS

### Study population

The RECOVER study is an observational prospective cohort study of HCWs with a history of PCR-confirmed SARS-CoV-2 infection. Eligible HCWs comprised any professional working in the Greater Montreal (Quebec, Canada) area healthcare facilities, including physicians, nurses, patient attendants, therapists, technicians, maintenance employees, food workers, administrative personnel, and researchers.

HCWs were recruited between August 17^th^, 2020, and April 8^th^, 2021, primarily through the McGill University Health Center (MUHC) / Centre hospitalier universitaire Sainte-Justine (CHUSJ) Vaccine Center. Additional recruitment took place at the Centre hospitalier de l’Université de Montréal (CHUM) and the Jewish General Hospital (JGH). Prospective participants were excluded if: (i) they were no longer working in a healthcare setting or had been furloughed as a preventive measure at the time of enrolment, (ii) they were not fluent in either French or English, (iii) they had no access to a cell phone or Internet for follow-up procedures, (iv) they were participating in a clinical trial for preventive treatment for COVID-19 and/or (v) they received a COVID-19 vaccine prior to enrolment. Planned follow-up period was at least 12 months for all participants and was extended to 18 months for participants that remained unvaccinated. A timeline of the study is presented in Figure 1.

**Figure 1:**
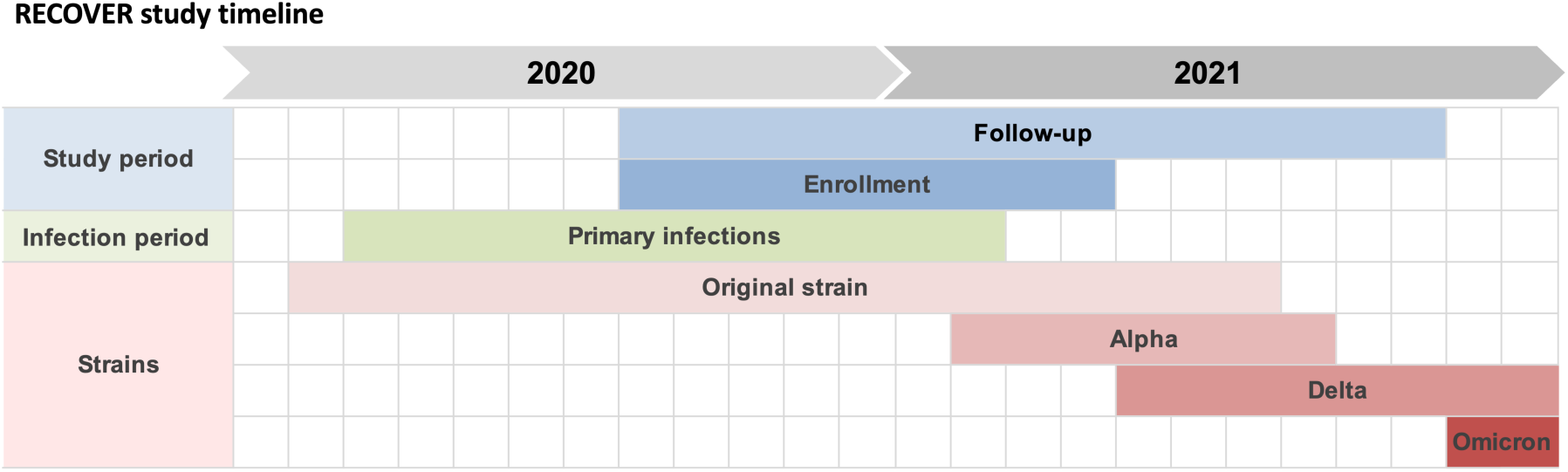
Timeline of the RECOVER study, overlayed with variants in circulation during the study period. Enrolment of participants was from August 17^th^ 2020 to April 8^th^ 2021. Follow-up period (for the results presented in this paper) was from August 17^th^ 2020 to October 19^th^ 2021. Enrolled participants acquired their primary infection between March 6^th^ 2020 and February 14^th^ 2021. Approximate periods for circulation of variants were derived from provincial surveillance data.

### Cohort size

Sample size was designed to measure a 3% risk of reinfection^1^ over 12 months with 95% confidence of a ±1.5% precision (precision equals half the estimated risk). Accounting for an estimated 10 to 15% loss to follow-up, a sample size of 570 HCWs was deemed optimal [18].

### Data collection, management, and analysis

#### Upon enrolment

Upon enrolment (day 0; D0), baseline demographic, clinical and biological data were obtained from each participant. Demographic data included date of birth, biological sex at birth, ethnicity, workplace, and profession. Clinical data included height, weight, medical history, medication, lifestyle information, and recent vaccination history. Detailed information about original COVID-19 illness was collected, including symptomatology, duration of symptoms, date of first positive PCR test, and need for hospitalization/advanced treatment. Blood samples were drawn at D0 for assessment of immune response to the original infection.

#### Follow-up

Every 2 weeks, an electronic questionnaire was sent to participants inquiring about new COVID-19 symptoms, need to consult a physician because of symptoms and history of recent significant exposure to a confirmed case of COVID-19. For that purpose, significant exposure was defined as at least 15 minutes within 2 meters of a confirmed infectious case without proper use of recommended personal protective equipment.

Quarterly in-person visits were planned at D90, D180, D270 and D360, where participants provided the following updates, if any: change in workplace, change in work duties, change in residual symptoms (if any were present at D0), new influenza-like illness, new medical condition, new medication, new vitamin intake and new vaccination. Additional blood samples for antibody serology and immune response assessment were drawn at quarterly visits.

Participants were asked to contact the research team between planned follow-ups if any of the following events occurred: (i) new symptoms onset, (ii) close contact with a confirmed case of COVID-19, (iii) significant exposure to a COVID-19 patient in the workplace without proper use of personal protective equipment or (iv) vaccination with any COVID-19 vaccine. If a participant reported new symptoms, a nasopharyngeal (NP) swab was performed. If the result was positive, an acute visit (2-4 days post symptoms onset) and convalescent visit (28-42 days post symptoms onset) were scheduled to obtain blood samples and details related to a possible reinfection (location/source of reinfection, symptomology, clinical course). For the first 30 participants^2^ who reported a significant exposure, a follow-up visit was scheduled four to seven days post exposure to collect an NP swab to ascertain asymptomatic reinfection.

Anonymized data was collected, stored, and managed using the secure web-based application REDCap [19]. Consent was obtained at enrolment and reviewed at each quarterly visit. Data was extracted from REDCap and exported into CSV/Microsoft Excel worksheets for initial management. Statistical analyses were performed with R version 4.1.2 and RStudio Version 2021.09.1. Kaplan-Meier survival analysis, Kaplan-Meier curves and Cox regression models were produced with the *survival* and *ggplot2* R packages. Tolerance threshold to type I error was defined as *α* = 0.05. Log-rank *p*-values derived from the score test were used for statistical comparison of Kaplan-Meier curves. Cox regressions modeled the hazard (probability per unit time) to obtain a negative serology test. Therefore, a hazard ratio (HR) higher than 1 represents a shorter average duration of seropositivity.

#### Outcomes

Primary outcomes were possible, probable, or confirmed reinfection with SARS-CoV-2, in absence of vaccination. Possible reinfection was defined as a positive PCR test less than 90 days after first infection. Probable reinfection was defined as a positive PCR test 90 days or more after the first positive test. Confirmed reinfection required either (i) evidence of infection by a known distinct variant or (ii) a variant that was not circulating at time of first infection or (iii) confirmation that the two strains were different by whole genome sequencing. Participants that received any vaccination against SARS-CoV-2 during follow-up were right-censored for reinfection outcomes at reception of the first dose. Our secondary outcome was serology status for IgG against the receptor-binding domain (RBD) of the SARS-CoV-2 spike as a function of time since primary infection, in participants that remained unvaccinated^3^.

#### Laboratory methods

Qualitative reverse-transcriptase PCR (RT-PCR) was performed on all samples. Upon reinfection, both the original strain and the reinfecting strain were sequenced, whenever possible, for phylogenetic studies to determine if the reinfecting and original strains differed. Original strains were obtained from the Laboratoire de santé publique du Québec (LSPQ) or from the laboratory where original testing was performed when unavailable from the LSPQ.

Antibody detection was performed on blood samples taken at D0 and at each quarterly visit. For symptomatic reinfections, acute and convalescent sera were collected for antibody testing. IgG levels were detected using an in-house, validated ELISA test based on the RBD of the spike protein. Validation of the ELISA was performed using a panel of 81 serum samples provided by the National Microbiology Laboratory of Canada; our RBD assay had a sensitivity of 95% and specificity of 100%. Participants were considered seropositive if the optical density (OD) was higher than the mean OD of negative controls^4^ plus three standard deviations.

## RESULTS

### Demographics and clinical data

The demographic characteristics of our cohort are presented in Table 1. Participants were in majority female (biological sex at birth; n=472, 83.0%) and Caucasian (n=451, 79.3%). Median age was 42 years (IQR 25-75: 32–50) and ranged from 18 to 75 years. Most HCWs worked in acute-care hospitals (n=308, 54.1%) or in public long-term care facilities (n=139, 24.4%). The most common reported profession was nurse/paramedic (n=230, 40.4%), followed by patient care attendant (n=73, 12.8%) and physician/medical resident (n=67, 11.8%).

**Table 1:** Demographic characteristics of the RECOVER cohort. The total number of enrolled HCWs is 569.

|  |  |  |
| --- | --- | --- |
| **Sex at birth** |  | **n (%)** |
|  | Female | 472 (83.0) |
|  | Male | 97 (17.0) |
| **Age** |  | **years** |
|  | Median (IQR) | 42 (18) |
|  | Range | 18 – 75 |
| **Ethnicity** |  | **n (%)** |
|  | Caucasian | 451 (79.3) |
|  | Middle-Eastern | 14 (2.5) |
|  | Latino | 22 (3.9) |
|  | Asian | 38 (6.7) |
|  | Black/African-American | 35 (6.2) |
|  | First Nations | 1 (0.2) |
|  | Other | 8 (1.4) |
| **Workplace** |  | **n (%)** |
|  | Hospital | 308 (54.1) |
|  | Public long-term care facility | 139 (24.4) |
|  | Community health center | 50 (8.8) |
|  | Private care facility | 12 (2.1) |
|  | Other | 60 (10.5) |
| **Staff group** |  | **n (%)** |
|  | Medical doctor/resident | 67 (11.8) |
|  | Nurse/paramedic | 230 (40.4) |
|  | Patient care attendant | 73 (12.8) |
|  | Therapist/other healthcare professional in regular contact with individual patients<sup>9</sup> | 84 (14.8) |
|  | Education/recreation | 12 (2.1) |
|  | Pharmacist/pharma assistant | 11 (1.9) |
|  | Technician | 16 (2.8) |
|  | Research staff | 7 (1.2) |
|  | Administration/management | 30 (5.3) |
|  | Maintenance/housekeeping | 15 (2.6) |
|  | Food service | 5 (0.9) |
|  | Other | 19 (3.3) |

<sup>9</sup> Includes: physiotherapists, occupational therapists, respiratory therapists, kinesiologists, nutritionists, social workers, psychologists, speech therapists, audiologists, electrophysiologists.

Baseline medical/lifestyle data are presented in Table 2. A total of 82 HCWs (14.4%) reported at least one medical condition considered a risk factor for severe COVID-19 illness per CDC guidelines [20]. Most HCWs were either overweight (25 ≤ BMI < 30 kg/m^2^; n=181, 31.8%) or obese (BMI ≥ 30 kg/m^2^; n=143, 25.1%), while only 8 HCWs were underweight (BMI < 18.5 kg/m^2^; 1.4%). Most participants (n=526, 92.4%) did not smoke tobacco products (including vaping products), cannabis or other drugs. Nearly a third of participants (n=182, 32.0%) reported regular vitamin D intake, with median weekly intake of 7000 IU.

**Table 2:** Risk factors for severe COVID-19 illness and lifestyle data.

| **Medical conditions associated with increased risk of severe COVID-19 illness<sup>10</sup>** | **n (%)** |
| --- | --- |
| Hypertension | 50 (8.8) |
| Chronic heart disease | 3 (0.5) |
| Diabetes | 21 (3.7) |
| Chronic lung disease<sup>11</sup> | 8 (1.4) |
| Chronic kidney disease | 1 (0.2) |
| Chronic liver disease | 7 (1.2) |
| Immune system suppression | 7 (1.2) |
| Other<sup>12</sup> | 10 (1.8) |
| At least 1 comorbidity | 82 (14.4) |
| **Body mass index** | **n (%)** |
| Underweight (BMI<18.5 kg/m<sup>2</sup>) | 8 (1.4) |
| Normal weight (18≤BMI<25 kg/m<sup>2</sup>) | 237 (41.7) |
| Overweight (25≤BMI<30 kg/m<sup>2</sup>) | 181 (31.8) |
| Obese (BMI≥30 kg/m<sup>2</sup>) | 143 (25.1) |
| **Lifestyle** | **n (%) or UI** |
| Smoking/vaping<sup>13</sup> | 43 (7.6) |
| Vitamin D regular intake | 182 (32.0) |
| Median weekly dose (IQR) | 7000 UI (3000) |

<sup>10</sup> There were no active cancers in our cohort, so this category is not included.
<sup>11</sup> Excluding asthma.
<sup>12</sup> Includes pregnancy, thalassemia, sickle cell anemia.
<sup>13</sup> Includes cannabis/cannabis products.

### Initial COVID-19 illness

Data regarding primary infections are reported in Table 3. In most participants (n=541, 95.1%), the primary infection resulted in symptomatic COVID-19 illness. The median duration of acute symptoms was 14 days (IQR 25-75: 7–21). Exposures leading to the primary infection occurred mostly in the workplace (n=425, 74.7%) or in the household (n=66, 11.6%), as reported by participants. Thirty-four HCWs (6.0%) required hospitalization to manage their illness, with 12 (2.1%) requiring oxygen therapy, two (0.4%) requiring intensive care and one (0.2%) requiring mechanical ventilation. Obesity was the only significant individual risk factor for hospitalization identified through multivariate logistic regression [adjusted OR=2.68 (1.11 – 6.72)]. Age, sex, ethnicity, pre-existing medical conditions other than obesity, vitamin D intake and smoking/vaping status were not significantly associated with hospitalization odds (see Supplementary Material for details). Median time between primary infection and enrolment was 177 days (IQR 25-75: 138–216).

**Table 3:** Description of the initial SARS-CoV-2 infections / COVID-19 illness episodes.

| **Symptomology** | **n (%)** |
| --- | --- |
| At least 1 symptom | 541 (95.1) |
| 8 symptoms or more | 314 (55.2) |
| No symptoms | 28 (4.9) |
| **Reported symptoms** | **n (%)<sup>14</sup>** |
| Fever | 288 (53.2) |
| Fatigue | 481 (88.9) |
| Myalgia | 365 (67.5) |
| Cough | 352 (65.1) |
| Sore throat | 262 (48.4) |
| Dyspnea | 240 (44.4) |
| Nasal congestion | 216 (39.9) |
| Anosmia | 394 (72.8) |
| Ageusia | 340 (62.8) |
| Chest pain | 183 (33.8) |
| Headache | 413 (76.3) |
| Dizziness | 174 (32.2) |
| Diarrhea | 190 (35.1) |
| Nausea | 145 (26.8) |
| Vomiting | 51 (9.4) |
| Abdominal pain | 92 (17.0) |
| Loss of appetite | 261 (48.2) |
| **Duration of acute symptoms, if present<sup>15</sup>** | **days** |
| Median | 14 |
| 25<sup>th</sup> percentile | 7 |
| 75<sup>th</sup> percentile | 21 |
| **Location of exposure** | **n (%)** |
| Occupational | 425 (74.7) |
| Household | 66 (11.6) |
| Other | 15 (2.6) |
| Unknown | 63 (11.1) |
| **Disease severity** | **n (%)** |
| Hospitalization | 34 (6.0) |
| Oxygen therapy | 12 (2.1) |
| Intensive care | 2 (0.4) |
| Mechanical ventilation | 1 (0.2) |

<sup>14</sup> Percentage among symptomatic individuals.<sup>15</sup> Two symptomatic individuals did not report duration of symptoms.

### Longitudinal follow-up

We detected 5 cases of probable reinfection between August 17^th^, 2020, and October 19^th^, 2021; their characteristics are reported in Table 4. We did not detect any possible nor confirmed reinfection. One participant reported a recurrence of acute symptoms 6 months after the first illness, but never tested positive again. Although this episode was suggestive of reinfection, it was excluded from analysis as it did not meet our primary outcome definitions. Cumulative time at risk for probable reinfection while unvaccinated amounted to 54 675 person-days, for an average reinfection incidence rate of 3.3 (1.1 – 7.8) per 100 person-year.

**Table 4:** Characteristics of reinfection cases. D0 = enrolment. PCR+ = positive PCR test. *Δt* = time interval. Ct = cycle threshold. First positive PCR refers to primary infection; second positive PCR refers to reinfection.

| Patient | Individual data | Case definition | $\Delta t$ D0 to 2<sup>nd</sup> PCR+ | $\Delta t$ 1<sup>st</sup> PCR+ to 2<sup>nd</sup> PCR+ | Ct 2<sup>nd</sup> PCR+ | 1<sup>st</sup> infection | 2<sup>nd</sup> infection | Last serology before 2<sup>nd</sup> PCR+ | Significant exposure within 14 days of 2<sup>nd</sup> PCR+ |
| --- | --- | --- | --- | --- | --- | --- | --- | --- | --- |
| 1 | Age range: 50-54<br>Work: Nurse | Probable reinfection | 50 days | 156 days | N/A | Symptomatic<br>Not hospitalized | Symptomatic<br>Not hospitalized | Positive | Yes |
| 2 | Age range: 55-59<br>Work: Physician | Probable reinfection | 7 days | 168 days | 34 | Symptomatic Hospitalized<br>(less than 1 day) | Asymptomatic | Positive | No |
| 3 | Age range: 40-44<br>Work: Nurse | Probable reinfection | 71 days | 94 days | 34 | Symptomatic Hospitalized<br>(less than 1 day) | Symptomatic<br>Not hospitalized | Positive | No |
| 4 | Age: 60-64<br>Work: Physician | Probable reinfection | 0 days<br>(detected at initial screening) | 257 days | 18 | Symptomatic<br>Not hospitalized | Asymptomatic | Positive | No |
| 5 | Age range: 30-34<br>Work: Nurse | Probable reinfection | 38 days | 104 days | 33 | Symptomatic<br>Not hospitalized | Asymptomatic | Negative | Yes |

Three out of five probable reinfections (60%) were asymptomatic, while only 5% of initial infections were asymptomatic (Fisher exact test p-value = 0.0013); no reinfections required hospitalization. One asymptomatic reinfection was detected through screening at enrolment, one was detected at the workplace and the third was detected through contact tracing. PCRs positive for reinfection generally reported low viral loads (Table 4). Significant exposure within two weeks before the 2^nd^ positive PCR was reported in three out of five (60%) reinfection cases.

The crude probability of remaining at risk for probable reinfection as a function of time since considered at risk is shown in Figure 2, panel A. Median time at risk of probable reinfection while unvaccinated was 86 days (95% CI: 80 – 93, IQR 25-75: 43 – 127). Twenty-nine participants did not contribute any person-time at risk, explaining the vertical drop of survival at D0: one participant was identified as reinfected through screening at D0 and the other 28 were vaccinated within 90 days from their first positive PCR. Overall, removal from the at-risk pool for probable reinfection while unvaccinated was mostly due to vaccination rather than reinfection itself.

**Figure 2:**
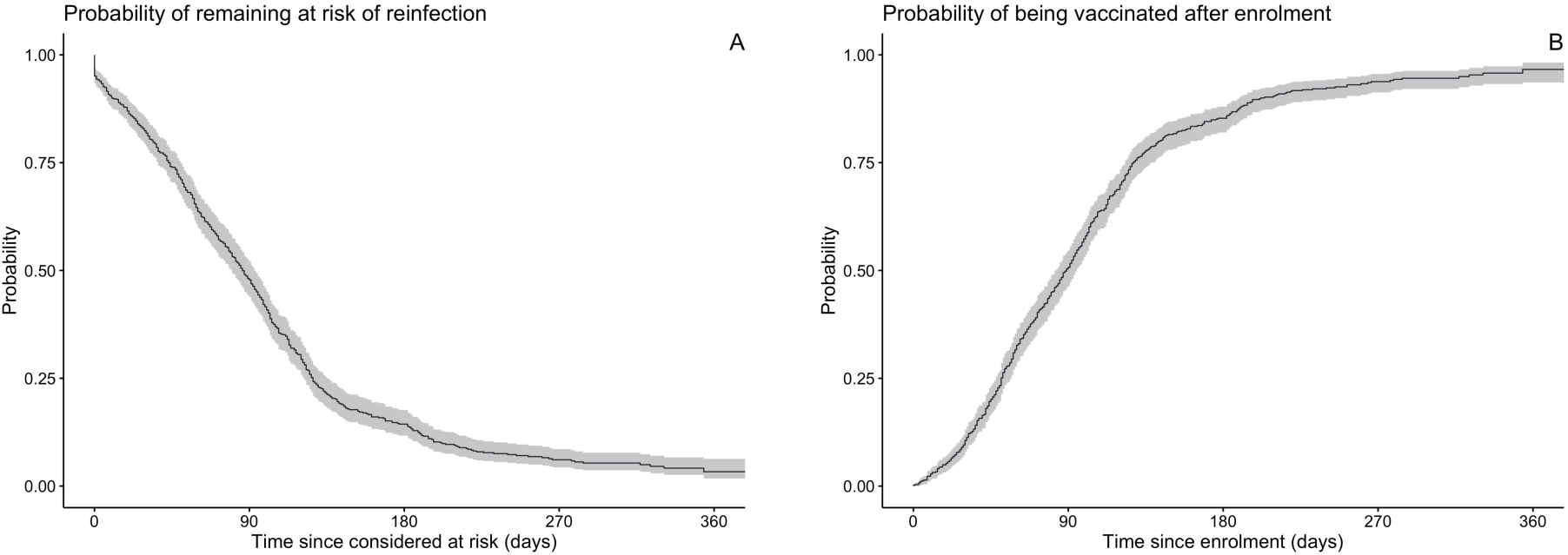
Longitudinal follow-up results for time at risk of primary outcome, displayed as Kaplan-Meier curves. Panel A: Probability of remaining in the pool of participants at risk of probable reinfection while unvaccinated, as a function of time since enrolment; event = reinfection or vaccination. Panel B: Probability of being vaccinated as function of time since enrolment; event = reception of a first dose of vaccine.

Seventy significant exposures to infectious COVID-19 cases, documented through bi-weekly questionnaires, occurred in 40 distinct HCWs during their at-risk period, for an incidence rate of 47 (36 – 59) significant exposures per 100 person-years. (Table 5).

**Table 5:** Demographic characteristics of the RECOVER participants with at least one significant exposure event while at risk of probable reinfection.

|  |  |  |
| --- | --- | --- |
| **Sex at birth** |  | **n (%)** |
|  | Female | 33 (82.5) |
|  | Male | 7 (17.5) |
| **Age** |  | **years** |
|  | Median (IQR) | 33 (13.5) |
|  | Range | 21 – 59 |
| **Ethnicity** |  | **n (%)** |
|  | Caucasian | 32 (80.0) |
|  | Other | 8 (20.0) |
| **Workplace** |  | **n (%)** |
|  | Hospital | 20 (50.0) |
|  | Public long-term care facility | 7 (17.5) |
|  | Community health center | 5 (12.5) |
|  | Private care facility | 1 (2.5) |
|  | Other | 7 (17.5) |
| **Staff group** |  | **n (%)** |
|  | Medical doctor/resident | 6 (15.0) |
|  | Nurse/paramedic | 18 (45.0) |
|  | Patient care attendant | 10 (25.0) |
|  | Therapist/other healthcare professional in regular contact with individual patients | 3 (7.5) |
|  | Other | 3 (7.5) |

Most participants (n=525, 92.3%) received at least 1 dose of vaccine against SARS-CoV-2 as of October 19^th^, 2021. Crude probability of having received at least 1 dose as function of time since D0 is shown in Figure 2, panel B. Among HCWs who received at least 1 dose, the median time between enrolment and reception of first dose was 89 days (95% CI: 82 – 95, IQR 25-75: 51 – 128). The probability of being vaccinated initially increases approximately linearly with time up until 125 days after D0, and then reached a plateau.

We observed a significant persistence of symptoms attributable to primary infection during follow-up. Figure 3 shows the crude Kaplan-Meier curve for the probability of presenting acute or residual symptoms as a function of time since infection; vaccinated individuals were right-censored for this analysis. More than half of participants reported residual symptoms 3 months after primary infection. The median time to complete resolution of symptoms was 164 days post-infection (95% CI: 90 – 241). The proportion of participants with residual symptoms declined with time, but about 30% still reported symptoms one year post-infection.

**Figure 3:**
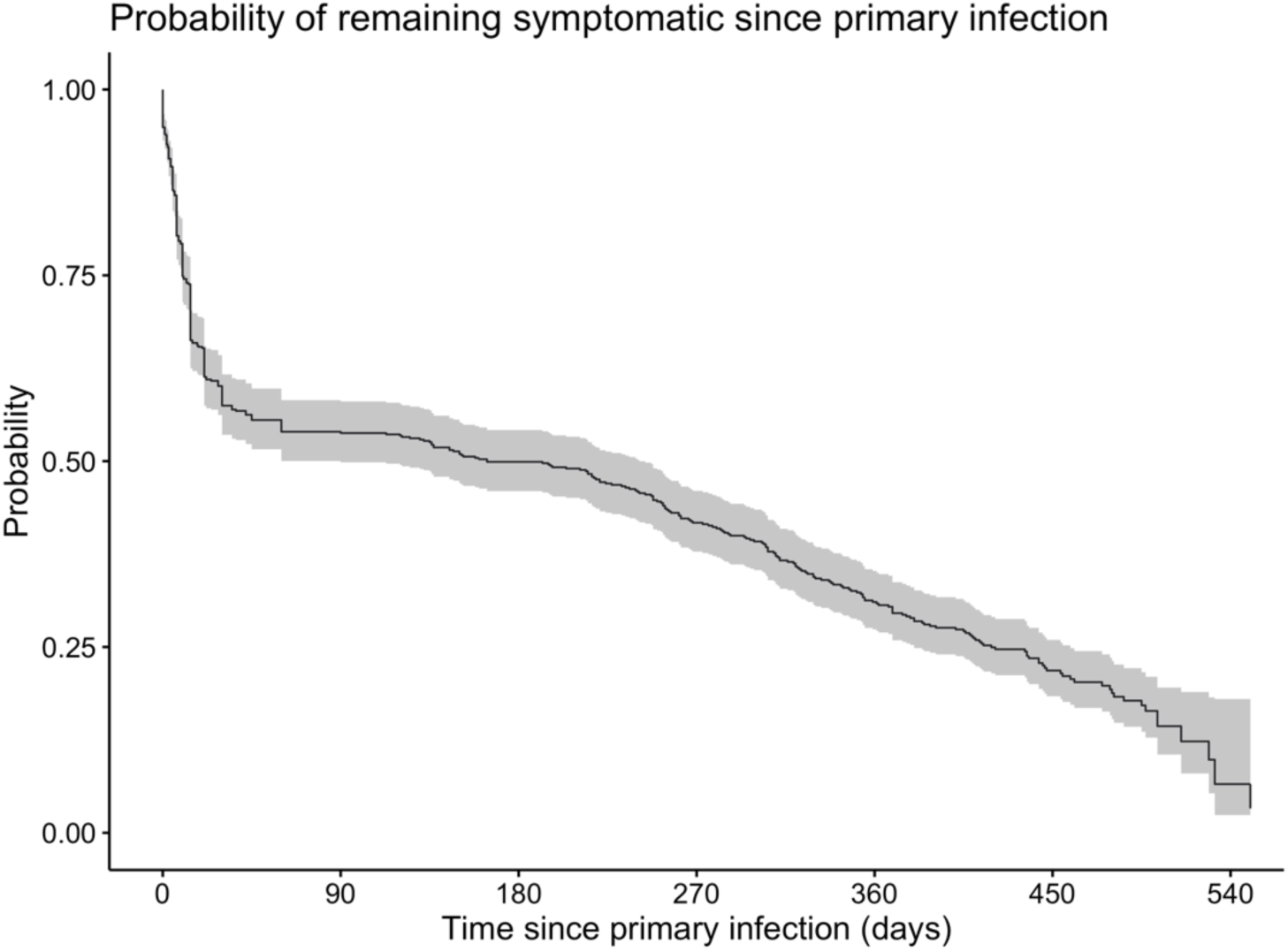
Kaplan-Meier curve for the probability of reporting symptoms attributable to the primary infection as a function of time since primary infection. For participants who reported no residual symptoms at enrolment (D0), the duration of symptoms was collected by questionnaire at D0. For participants who reported residual symptoms at D0, we considered them as symptomatic until they declared complete resolution of symptoms at a quarterly visit.

At time of writing, serology results were fully available for D0, D90 and D180 visits. We performed time-to-event Kaplan-Meier analysis on our serology data, where the event was a negative serology. Crude probability of remaining seropositive as a function of time since primary infection and in the absence of vaccination is shown in Figure 4, panel A. One hundred sixty-three (163) participants had a negative serology either at D0 (n=140, 24.6%) or during follow-up (n=23, 4.0%). Median duration of seropositivity was 415 days since primary infection (95% CI: 406 – Infinity)^5^. Kaplan-Meier curves stratified by presence/absence of symptoms at primary infection are shown in Figure 4, panel B. We observed that prior to vaccination, individuals with symptomatic primary infection remained seropositive longer than individuals with asymptomatic primary infection (4.9% were asymptomatic; Table 3). The median duration of seropositivity was 420 days (95% CI: 406 – Infinity) in initially symptomatic participants, compared to 213 days (95% CI: 161 – Infinity) in initially asymptomatic participants (p<0.0001). Kaplan-Meier curves further stratified according to number of symptoms are shown in Figure 4, panel C; mean number of self-reported symptoms was between 7 and 8. Individuals with 8 symptoms or more, defined as polysymptomatic, remained seropositive longer than individuals with 1 to 7 symptoms, defined as paucisymptomatic (p<0.0001). The median duration of seropositivity in the polysymptomatic category was undefined^6^, while median duration of seropositivity in paucisymptomatic category was 420 days (95% CI: 329 – Infinity). Finally, Kaplan-Meier curves stratified by ethnicity are shown in Figure 4, panel D. We observed that non-Caucasian participants remained seropositive significantly longer than Caucasians (p = 0.0031). Median duration of seropositivity in non-Caucasians was undefined^7^, while median duration of seropositivity in Caucasians was 406 days (95% CI: 386 – Infinity).

**Figure 4:**
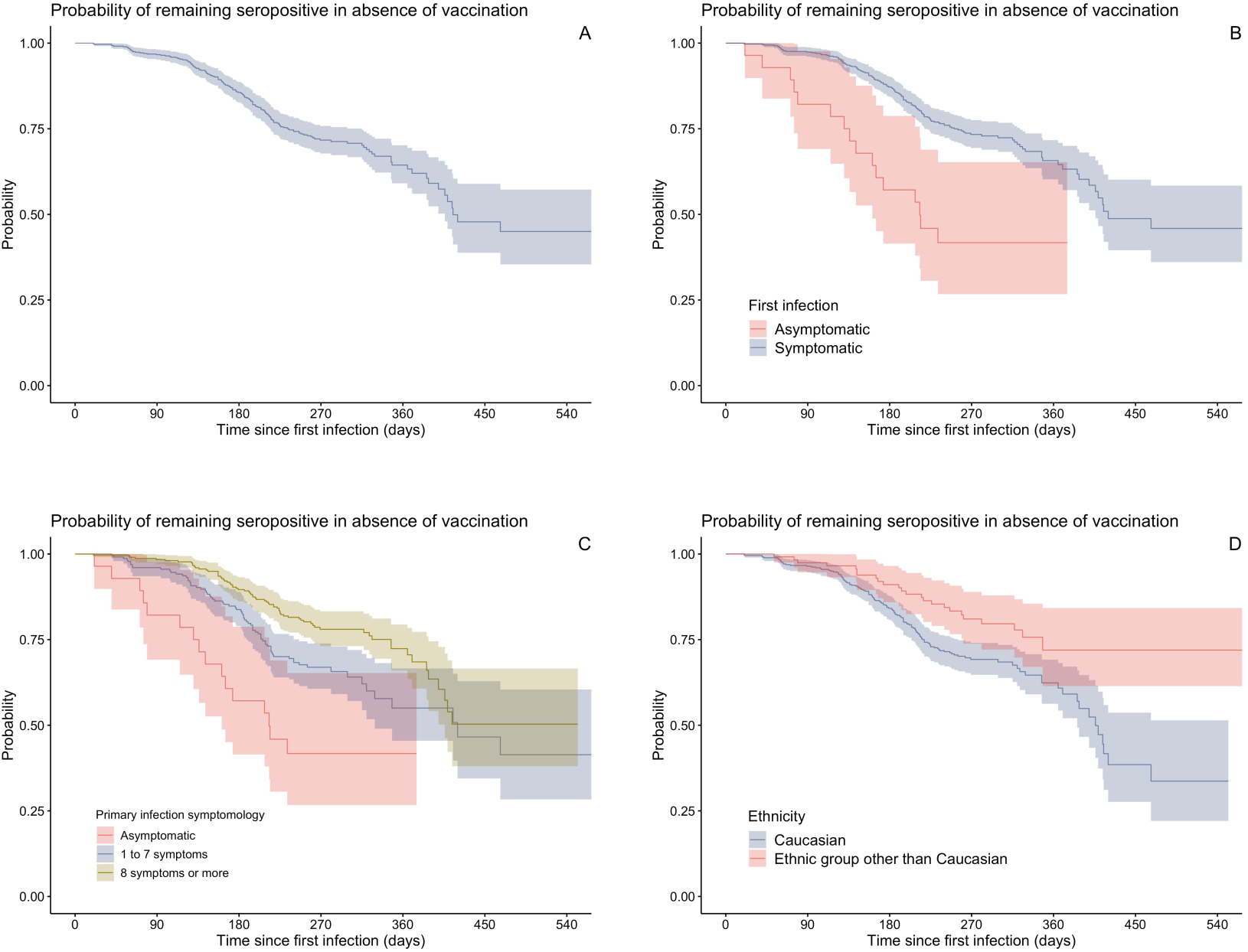
Longitudinal follow-up results for serology, displayed as Kaplan-Meier curves. All curves represent the probability of remaining seropositive as function of time since primary infection, in absence of vaccination. Negative serology was the primary event and vaccination was a censoring event. Panel A: Crude Kaplan-Meier curve including all participants. Panel B: Kaplan-Meier curves stratified by symptomatology of primary infection alone (asymptomatic vs symptomatic). Panel C: Kaplan-Meier curves stratified by ethnic group alone (Caucasian vs ethnic group other than Caucasian). Panel D: Kaplan-Meier curves stratified by symptomology of first infection and ethnic group; confidence intervals not shown for clarity.

We performed a multivariate Cox regression on our serology data to adjust for potential confounders/effect modifiers. Participants with asymptomatic primary infection were less likely to remain seropositive over time [adjusted HR for testing seronegative=2.25 (1.30 – 3.91)] when compared to participants with paucisymptomatic primary infection. Conversely, participants with polysymptomatic primary infection were more likely to remain seropositive [adjusted HR=0.65 (0.46 – 0.91)] when compared to paucisymptomatics. Participants aged 55 years and over were more likely to remain seropositive compared to younger participants [adjusted HR=0.52 (0.30 – 0.92)]. Obese participants were more likely to remain seropositive compared to participants of normal weight [adjusted HR=0.44 (0.27 – 0.72)]. The hazard ratio for testing seronegative did not significantly differ from the null when comparing overweight or underweight participants to normal weight participants. Lastly, non-Caucasian HCWs were more likely to remain seropositive compared to Caucasian participants [adjusted HR=0.48 (0.31 – 0.75)]. Hospitalization, duration of symptoms, comorbidities other than obesity, sex, smoking/vaping, vitamin D intake, worksite, profession, and household size were not significantly associated with an increased or decreased hazard of testing seronegative (see Supplementary Data for details).

Fifty-six participants (9.8%) were either lost to follow-up or withdrew from the study at the time of interim analysis.

## DISCUSSION

We described the interim results of the RECOVER study over the first 14 months of follow-up. Our study shows that reinfection in unvaccinated HCWs with a history of PCR-confirmed SARS-CoV-2 infection remains a rare event over the first several months post-infection. Our measured reinfection incidence rate of 3.3 per 100 person-year is generally concordant with rates observed by other authors in HCWs and the general population. By comparison, Gallais et al. observed a reinfection incidence rate of 0.40 per 100 person-years in a cohort of French HCWs over 13 months [21]. Hall et al. followed a cohort of English HCWs prospectively for one year (SIREN study) and observed a reinfection incidence rate of 2.8 per 100 person-year [7]. Another English cohort study in HCWs by Lumley et al. reported a reinfection incidence rate of 0.47 per 100 person-years [9]. Other studies investigated reinfection rates in the general population. Abu-Raddad et al. estimated the reinfection incidence rate in Qatar at 1.3 per 100 person-years, using a cohort of laboratory-confirmed primary infections [22]. In a Danish population-level observational study by Hansen et al., the reinfection incidence rate was estimated at 2.0 per 100 person-years [12]. Other studies reported risk of reinfection in various cohorts [6,23–25], but did not report incidence rates^8^. It is important to note that the reinfection rates observed in these studies predate the Omicron wave and depend on the incidence rates in the general population of each region/country, which limits inter-study comparability.

Our measured incidence rate of self-reported significant exposure was approximately 14 times higher than our reinfection incidence rate. However, it remains unclear if this rate ratio constitutes a reliable measure of protection because: (i) we did not have a cohort naïve to SARS-CoV-2 to establish baseline comparison values for infection and significant exposure incidence rates and (ii) it is likely that many significant exposure events were either unreported or unrecognized by participants.

Our Kaplan-Meier analysis shows that the overall probability of remaining seropositive up to 300 days after the initial infection is approximately 70% in the absence of vaccination. Duration of seropositivity was significantly positively correlated with the number of symptoms at primary infection; duration of seropositivity was longest in polysymptomatics and shortest in asymptomatics. Duration of seropositivity was significantly longer in non-Caucasian participants compared to Caucasian participants. Although not explained in our regression models, we hypothesize that the effect of ethnicity on duration of seropositivity could be attributed to profession, workplace and household size [26,27], which could lead to more re-exposures without symptomatic disease that thus went undetected. Finally, it remains uncertain whether positive serology constitutes a strong correlate of protection [28]. However, emerging evidence indicates that higher levels of IgG seem correlated with higher neutralization capacities [7,12,29].

Our epidemiological and serological evidence supports the hypothesis that primary infection by SARS-CoV-2 confers significant protection against reinfection for at least several months, and up to the spread of the Omicron variant.

Our study has several strengths. First, our prospective cohort is representative of the population of HCWs in the Greater Montreal area, through our multicentric recruitment process and permissive eligibility criteria. Participants were closely monitored during follow-up, with frequent electronic questionnaires and in-person visits, which decreases recall/memory bias. Data collection was exhaustive and included a wealth of demographic, clinical, and biological information, allowing adjustment/stratification for many potential confounders and effect measure modifiers. HCWs constituted an ideal population for studying reinfection risk and protection conferred by natural infection because they are more frequently exposed to SARS-CoV-2 than the general population, increasing the validity of our measurements. Finally, most participants remained enrolled in the study, limiting selection bias from loss to follow-up.

Our study nevertheless has some limitations. First, we observed a very small number of reinfections, hence limiting statistical power. Specifically, we could not determine whether demographic and/or clinical individual characteristics were associated with an increased or decreased reinfection incidence rate. It is also likely that we missed cases of asymptomatic reinfections, since we did not systematically screen all participants with NP swabs on a regular basis (e.g., every 2 weeks). Participants were randomly swabbed only twice during the entire follow-up, which could allow for asymptomatic reinfections to remain undetected. Our measured reinfection incidence rate thus probably underestimates the true reinfection rate, especially if asymptomatic reinfections are PCR-positive for only a short time. Our study could not identify any case of confirmed reinfection by genetic comparison of viral strains, because material recovered in reinfection swabs was insufficient to allow genome sequencing. These low viral loads suggest that reinfected individuals may be less likely to transmit SARS-CoV-2 to susceptible individuals compared to naïve individuals who become infected [30], with the caveat that our study was conducted prior to the spread of the Omicron variant.

Vaccines against SARS-CoV-2 became available to participants about 4 months after the initiation of our study. Since participant recruitment was not yet completed when vaccination started, time between enrolment and vaccination was highly heterogeneous, which could impart selection bias. Therefore, we could not calculate a meaningful risk of reinfection in previously infected but unvaccinated individuals over 12 months.

Finally, our cohort is composed mostly of young, female, Caucasian and generally healthy HCWs. This limits the generalizability of our results to other populations, and our capacity to reveal and/or explain possible associations between covariates like sex, age, ethnicity and presence of comorbidities, and our outcomes of reinfection and loss of measurable serum IgG.

## SUMMARY

Reinfection by SARS-CoV-2 remained a rare event several months after first infection, among a population of 569 Canadian healthcare workers. Reinfection episodes were milder than original illness and were characterized by low viral loads. Initial infection induced detectable serum IgG levels in the majority (75%) of participants at enrolment. Duration of seropositivity was positively correlated to the number of symptoms during the acute phase of primary infection, age over 55 years, obesity and non-Caucasian ethnicity. Our study provides epidemiological and serological evidence that initial infection by SARS-CoV-2 confers protection against reinfection for several months. Additional research is needed to assess the frequency of asymptomatic reinfections and their relative transmissibility compared to primary infections. There is also a need to complement these findings with an in-depth analysis of the humoral and cellular responses to SARS-CoV-2 infection.

## Data Availability

All data produced in the present study are available upon reasonable request to the authors

## Footnotes

1 At study design, estimates of reinfection risk were obtained from literature on other human coronaviruses, which suggested a higher risk of reinfection than what has since been observed with SARS-CoV-2. Vaccines were being developed and were not available at the time of study design.

2 This number was limited to 30 because of budget constraints.

3 In Quebec, there was no mandatory vaccination for HCWs to continue working.

4 Controls were negative sera obtained in the pre-pandemic era.

5 Assuming all participants were seropositive at time of primary infection.

6 Because less than 50% of polysymptomatics became seronegative during follow-up.

7 Because less than 50% of non-Caucasians became seronegative during follow-up.

8 Peer-reviewed reinfection incidence rates cited in this section were estimated before the spread of the omicron variant; they may not reflect reinfection incidence rates during the first omicron wave.

## REFERENCES

1. WHO Coronavirus (COVID-19) Dashboard. Available at: https://covid19.who.int. Accessed 24 January 2022.

2. Nicola M, Alsafi Z, Sohrabi C, et al. The socio-economic implications of the coronavirus pandemic (COVID-19): A review. Int J Surg Lond Engl 2020; 78:185– 193.

3. Burki TK. Omicron variant and booster COVID-19 vaccines. Lancet Respir Med 2021; 0. Available at: https://www.thelancet.com/journals/lanres/article/PIIS2213-2600(21)00559-2/fulltext?s=09. Accessed 28 December 2021.

4. Sawal I, Ahmad S, Tariq W, Tahir MJ, Essar MY, Ahmed A. Unequal distribution of COVID-19 vaccine: A looming crisis. J Med Virol 2021; 93:5228–5230.

5. Cardenas NC. ‘Europe and United States vaccine hesitancy’: leveraging strategic policy for ‘Infodemic’ on COVID-19 vaccines. J Public Health Oxf Engl 2021;

6. Dimeglio C, Herin F, Miedougé M, Martin-Blondel G, Soulat JM, Izopet J. Protection of healthcare workers against SARS-CoV-2 reinfection. Clin Infect Dis 2021;

7. Hall VJ, Foulkes S, Charlett A, et al. SARS-CoV-2 infection rates of antibody-positive compared with antibody-negative health-care workers in England: a large, multicentre, prospective cohort study (SIREN). Lancet 2021; 397:1459–1469.

8. Hanrath AT, Payne BAI, Duncan CJA. Prior SARS-CoV-2 infection is associated with protection against symptomatic reinfection. J Infect 2021; 82:e29–e30.

9. Lumley SF, O’Donnell D, Stoesser NE, et al. Antibody Status and Incidence of SARS-CoV-2 Infection in Health Care Workers. N Engl J Med 2021; 384:533–540.

10. Abu-Raddad LJ, Chemaitelly H, Coyle P, et al. SARS-CoV-2 antibody-positivity protects against reinfection for at least seven months with 95% efficacy. EClinicalMedicine 2021; 35:100861.

11. Cohen C, Kleynhans J, von Gottberg A, et al. SARS-CoV-2 incidence, transmission and reinfection in a rural and an urban setting: results of the PHIRST-C cohort study, South Africa, 2020-2021. medRxiv 2021; :2021.07.20.21260855.

12. Hansen CH, Michlmayr D, Gubbels SM, Mølbak K, Ethelberg S. Assessment of protection against reinfection with SARS-CoV-2 among 4 million PCR-tested individuals in Denmark in 2020: a population-level observational study. Lancet 2021; 397:1204–1212.

13. Pilz S, Chakeri A, Ioannidis JP, et al. SARS-CoV-2 re-infection risk in Austria. Eur J Clin Invest 2021; :e13520.

14. Rennert L, McMahan C. Risk of SARS-CoV-2 reinfection in a university student population. Clin Infect Dis 2021;

15. Sheehan MM, Reddy AJ, Rothberg MB. Reinfection Rates among Patients who Previously Tested Positive for COVID-19: a Retrospective Cohort Study. Clin Infect Dis 2021;

16. Vitale J, Mumoli N, Clerici P, et al. Assessment of SARS-CoV-2 Reinfection 1 Year After Primary Infection in a Population in Lombardy, Italy. JAMA Intern Med 2021;

17. Pulliam JRC, Schalkwyk C van, Govender N, et al. Increased risk of SARS-CoV-2 reinfection associated with emergence of the Omicron variant in South Africa. 2021: 2021.11.11.21266068. Available at: https://www.medrxiv.org/content/10.1101/2021.11.11.21266068v2. Accessed 28 December 2021.

18. Naing L, Winn T, Nordin R. Pratical Issues in Calculating the Sample Size for Prevalence Studies. Arch Orofac Sci 2006; 1.

19. Harris PA, Taylor R, Thielke R, Payne J, Gonzalez N, Conde JG. Research electronic data capture (REDCap)--a metadata-driven methodology and workflow process for providing translational research informatics support. J Biomed Inform 2009; 42:377–381.

20. CDC. COVID-19 and Your Health. 2020. Available at: https://www.cdc.gov/coronavirus/2019-ncov/need-extra-precautions/people-with-medical-conditions.html. Accessed 27 September 2021.

21. Gallais F, Gantner P, Bruel T, et al. Evolution of antibody responses up to 13 months after SARS-CoV-2 infection and risk of reinfection. EBioMedicine 2021; 71:103561.

22. Abu-Raddad LJ, Chemaitelly H, Malek JA, et al. Assessment of the Risk of Severe Acute Respiratory Syndrome Coronavirus 2 (SARS-CoV-2) Reinfection in an Intense Reexposure Setting. Clin Infect Dis Off Publ Infect Dis Soc Am 2020; 73:e1830–e1840.

23. Graham MS, Sudre CH, May A, et al. Changes in symptomatology, reinfection, and transmissibility associated with the SARS-CoV-2 variant B.1.1.7: an ecological study. Lancet Public Health 2021; 6:e335–e345.

24. Brouqui P, Colson P, Melenotte C, et al. COVID-19 re-infection. Eur J Clin Invest 2021; :e13537.

25. Bongiovanni M, Marra AM, Bini F, Bodini BD, Carlo DD, Giuliani G. COVID-19 reinfection in healthcare workers: a case series. J Infect 2021; Available at: https://www.journalofinfection.com/article/S0163-4453(21)00165-1/pdf.

26. Government of Canada SC. Montréal – A data story on Ethnocultural Diversity and Inclusion in Canada. 2019. Available at: https://www150.statcan.gc.ca/n1/pub/11-631-x/11-631-x2019001-eng.htm. Accessed 25 January 2022.

27. Government of Canada SC. The contribution of immigrants and population groups designated as visible minorities to nurse aide, orderly and patient service associate occupations. 2020. Available at: https://www150.statcan.gc.ca/n1/pub/45-28-0001/2020001/article/00036-eng.htm. Accessed 25 January 2022.

28. Krammer F. A correlate of protection for SARS-CoV-2 vaccines is urgently needed. Nat Med 2021; 27:1147–1148.

29. Krammer F. Correlates of protection from SARS-CoV-2 infection. The Lancet 2021; 397:1421–1423.

30. Iwasaki A. What reinfections mean for COVID-19. Lancet Infect Dis 2021; 21:3–5.

