## Supplemental Tables 1 and 2 for "The REinfection in COVID-19 Estimation of Risk (RECOVER) study: Reinfection and serology dynamics in a cohort of Canadian healthcare workers"

SUPPLEMENTARY MATERIAL

*Logistic regression model for hospitalization risk factors*

We investigated whether known risk factors for severe COVID-19 were significant predictors of hospitalization risk in our cohort. We used odds ratios derived from penalized maximum likelihood (PLM) logistic regression (R package *logistf*) to estimate the association between potential risk factors and probability of hospitalization. We chose this approach since the outcome is binary, has relatively low prevalence (n=34, 6.0%) and cell counts in some categories are low. Regression results for the base model are shown in Table S1.

**Table S1** : Multivariate logistic regression model for the odds of hospitalization as function of known severe COVID-19 risk factors.

| Variable | Odds ratio of hospitalization | 95% CI | *p*-value |
|---|---:|---:|---:|
| Sex | 1.42 | 0.59 – 3.15 | 0.420 |
| Age over 55 years | 2.39 | 0.99 – 5.47 | 0.054 |
| Ethnicity | 1.26 | 0.53 – 2.78 | 0.584 |
| **Obesity** | **2.68** | **1.11 – 6.72** | **0.029** |
| Overweight | 1.20 | 0.46 – 3.07 | 0.707 |
| Underweight | 1.57 | 0.012 – 14.8 | 0.757 |
| Comorbidity | 1.18 | 0.44 – 2.90 | 0.724 |
| Smoking/vaping | 0.16 | 0.001 – 1.20 | 0.085 |

Model results did not vary significantly when changing the age cutoff defining the older age category. The baseline model with cutoff at 55 years of age has the lowest AIC value and has thus been retained as the best fit model. Obesity was the only covariate significantly associated with higher odds of hospitalization.

*Cox regression model for serology time series data*

We built exploratory Cox regression models to investigate which individual factors act as predictors of persistence of IgG seropositivity after primary infection. The event of interest for this analysis was a negative serology. Regression models were built sequentially with the *coxph*() function to identify candidate predictors and potential confounders. Results for the coefficients of the final model are shown in Table S2 below. The proportional hazard hypothesis was tested with the *cox.zph*() function. The p-value for the global hypothesis test was 0.065; the p-values for individual covariates were all above the 0.05 threshold. The serology data is thus compatible with proportional hazards at the 95% confidence level.

Further adjustment for smoking/vaping, vitamin D intake, workplace, profession, and household size did not significantly change model results.

**Table S2** : Multivariate Cox regression model for the hazard of testing seronegative

| Variable | Hazard ratio | 95% CI | *p*-value |
|---|---|---|---|
| **Asymptomatic primary infection** | **2.25** | **1.30 – 3.91** | **0.004** |
| **Polysymptomatic primary infection** | **0.65** | **0.46 – 0.91** | **0.012** |
| Hospitalization | 0.37 | 0.12 – 1.19 | 0.095 |
| Sex | 1.02 | 0.65 – 1.60 | 0.916 |
| **Age over 55 years** | **0.52** | **0.30 – 0.92** | **0.025** |
| Any comorbidity | 1.13 | 0.65 – 1.98 | 0.664 |
| **Non-Caucasian** | **0.48** | **0.31 – 0.75** | **0.001** |
| **Obesity** | **0.44** | **0.27 – 0.72** | **0.001** |
| Overweight | 0.82 | 0.58 – 1.17 | 0.270 |
| Underweight | 1.24 | 0.39 – 4.00 | 0.716 |
